# Sleep-related traits and attention-deficit/hyperactivity disorder comorbidity: shared genetic risk factors, molecular mechanisms, and causal effects

**DOI:** 10.1101/2020.09.13.20193813

**Authors:** Marina Xavier Carpena, Carolina Bonilla, Alicia Matijasevich, Thais Martins-Silva, Julia P. Genro, Mara Helena Hutz, Luis Augusto Rohde, Luciana Tovo-Rodrigues

**Affiliations:** Post-Graduate Program in Epidemiology, Federal University of Pelotas, Pelotas, Brazil; Department of Preventive Medicine, Faculty of Medicine, University of São Paulo, São Paulo, Brazil; Population Health Sciences, University of Bristol, Bristol, United Kingdom; Programa de Pós-Graduação em Biociências, Universidade Federal de Ciências da Saúde de Porto Alegre, Porto Alegre, Brazil; Federal University of Rio Grande do Sul, Department of Genetics, Porto Alegre, Brazil; Federal University of Rio Grande do Sul, Department of Psychiatry, Child & Adolescent Psychiatry Unit,Hospital de Clinicas de Porto Alegre, Porto Alegre, Brazil

**Keywords:** ADHD, genetics, sleep, circadian rhythm, neurodevelopment, Mendelian randomization

## Abstract

**Study Objectives:** To evaluate the level of shared genetic components between attention-deficit/hyperactivity disorder (ADHD) and sleep phenotype, common pathways between them and a possible causal relationship between traits.

**Methods:** We used summary statistics of the largest genome-wide association studies available for ADHD and sleep-related phenotypes including insomnia, napping, daytime dozing, snoring, ease getting up, daytime sleepiness, sleep duration and chronotype. We estimated the genomic correlation between ADHD and sleep-related traits using cross-trait LD-score regression and investigated potential common mechanisms using gene-based cross-trait metanalyses and functional enrichment analyses. The causal effect between the sleep related traits and ADHD was estimated with two sample Mendelian randomization (TSMR), using the Inverse Variance Weighted method as the main estimator.

**Results:** Positive genomic correlation between insomnia, daytime napping, daytime dozing, snoring, daytime sleepiness, short and long sleep duration, and ADHD were observed. Insomnia, sleep duration, daytime sleepiness, and snoring shared genes with ADHD, which were involved in neurobiological functions and regulatory signaling pathways. The TSMR approach supported a causal effect of insomnia, daytime napping, and short sleep duration on ADHD, and of ADHD on long sleep duration and chronotype.

**Conclusion:** Our findings suggest that the comorbidity between sleep phenotypes and ADHD may be mediated by common genetic factors with an important role on neuronal signaling pathways. In addition, it may also exist a causal effect of sleep disturbances and short sleep duration on ADHD, reinforcing the role of these sleep phenotypes as predictors or early markers of ADHD.

**Statement of Significance:** Our findings suggest that the comorbidity between sleep phenotypes and ADHD may be linked to common genetic factors with an important role on neuronal signaling pathways. They also show that a causal effect of sleep disturbances and short sleep duration on ADHD may exist, reinforcing the role of these sleep phenotypes as ADHD early markers, being able to predict the disorder. This study adds important information about the relationship between sleep, circadian rhythm, and ADHD since it raises new questions about the complexity of the phenomenon involving them and may also provide new treatment insights in this research field.

## Introduction

Attention-deficit/hyperactivity disorder (ADHD) is one of the most common and highly heritable neurodevelopment disorders worldwide ^1,2^. Sleep disturbances and circadian disruption are common comorbid medical conditions that have been receiving growing attention in recent investigations. Several studies have shown that individuals affected by ADHD present higher prevalence of sleep and circadian disruptions than controls ^3,4^. In childhood, poor sleep quality is a frequent trait, being present in about 55–75% of children with ADHD ^5-7^. In adults sleep problems are also significantly associated with ADHD, being present in 60-80% of adult individuals with ADHD ^8-10^. Also, delayed sleep phase disorder, a circadian rhythm sleep-wake disorder, affects 73-78% of patients with ADHD in both childhood and adult life ^3,11,12^

Although the relationship between ADHD and sleep-related phenotypes may emerge from a complex environmental and genetic context, the potential mechanisms underlying the comorbidity, as well as causal relationships, are still not completely understood. Core clock gene impairment may link both conditions. In mice, modulation of the expression of the *Clock Per1/2* and *c-Fos* genes in the brain while under methylphenidate and atomoxetine treatments was observed ^13-15^, and in zebrafish, the *perlb-/-* mutation was able to reproduce an ADHD mimic behavior ^16^. The alteration of the expression of these genes led to changes in norepinephrine and dopamine regulation, which are important neurotransmitters for both sleep regulation and ADHD symptomatology ^17-19^. In humans, expression and candidate-gene studies have supported a role of the *CLOCK, PER2, BMAL1* and *CSNK1E* genes as potential links between sleep/circadian traits and ADHD ^20-25^. However, to the best of our knowledge, a broader investigation of shared genetics from a genomic perspective has not been performed to date.

The direction of the association and whether there is a causal effect are unanswered questions and hot topics of a current debate in the field ^26^. Sleep deprivation and sleep problems are known to mimic or aggravate ADHD symptoms, and to regulate emotion, cognition and behavior ^3,4^. On the other hand, ADHD-like behaviors may also lead to altered patterns of sleep ^26,27^. Early sleep disturbances predicting ADHD in the future are supported by longitudinal evidence ^28-32^. In addition, the opposite direction, in which ADHD may precede sleep disturbances and sleep deprivation is also reported in the literature ^33-35^. Thus, studies exploring causal relationships with different designs may contribute important evidence to better understand this relationship.

Evidence about genetic etiologies for ADHD and sleep related traits has considerably increased in the last five years. Recent genome wide association studies (GWAS) have reported novel genes associated with sleep and circadian phenotypes, besides some already uncovered via candidate gene studies or animal models. Therefore, we were able to identify genome-wide markers associated with insomnia^36,37^ daytime dozing^36^, daytime napping^36^, snoring^36^, ease getting up ^36^, daytime sleepiness ^38^, chronotype ^36,39^ and sleep duration ^36,40,41^. Likewise, a powerful GWAS study was recently published, describing the first top-hits for the ADHD disorder ^42^.

The recent publication of GWAS studies for sleep, circadian and ADHD phenotypes allows for the understanding of potential shared genetic and molecular mechanisms between sleep disturbances and ADHD, the dissection of possible differences in the association between specific sleep traits and ADHD, as well as the investigation on causality and the direction of this association. In this study we used the genomic data of the largest GWAS to date of several sleep-related phenotypes (sleep disturbances, sleep duration and chronotype) and ADHD to evaluate the level of shared genetic factors between traits, to examine possible common pathways between them and to explore a possible causal relationship between the traits through a bidirectional two sample Mendelian randomization approach (TSMR).

## Methods

### Datasets

In this study, we have included the data (summary statistics) of the largest GWAS published to date. These studies had been approved by local ethics committees and had obtained the required informed consents (detailed in previously published papers)^36-41,43,44^. The overall study design is shown in Figure 1.

**Figure 1.**
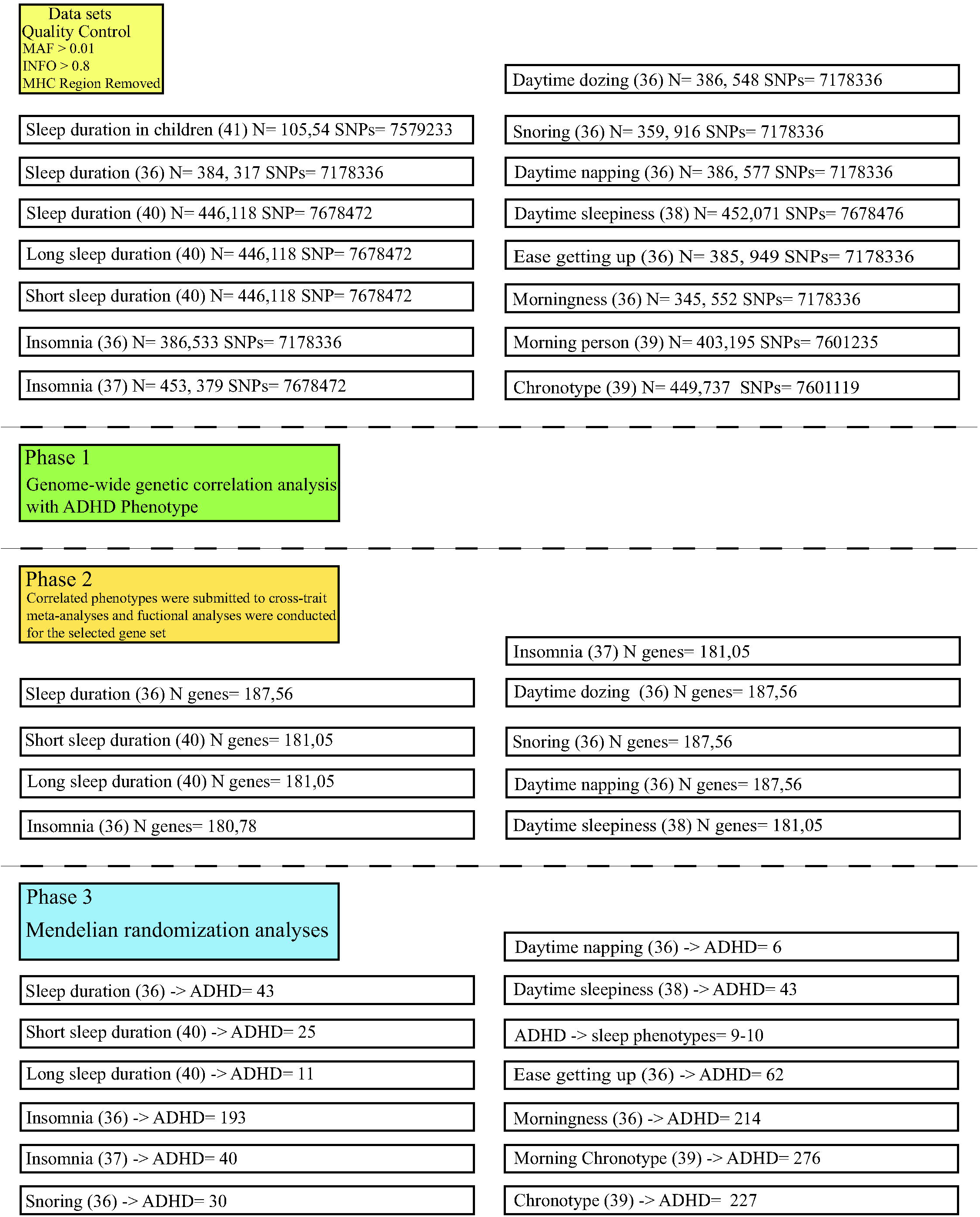
Overall study design and datasets illustration

#### ADHD phenotype

We used data from 20,183 ADHD cases and 35,191 controls in total, which included children and adults from the iPSYCH study and the 11 European studies belonging to the Psychiatric Genomic Consortium (PGC) ^42^. A genome-wide association analysis was conducted in each cohort using logistic regression, assuming additive genetic effects. Summary statistics of the results of the ADHD GWAs meta-analysis of iPSYCH and the PGC samples are available on their websites (https://www.med.unc.edu/pgc/results-and-downloads and http://ipsych.au.dk/downloads/).

#### Sleep-related phenotypes

We used the most recent publicly available summary association results for chronotype^36,39^, sleep duration ^36,40^, insomnia ^36,37^, daytime sleepiness ^38^, daytime napping ^36^, daytime dozing ^36^, ease getting up ^36^ and snoring ^36^ available on the UK Biobank (UKBB) website (https://www.ukbiobank.ac.uk/tag/gwas/) (Table S1). UKBB is a national prospective cohort that recruited more than 500,000 men and women from across the United Kingdom, aged 40-69 years between 2006 and 2010. For the chronotype analysis, 449,737 individuals with chronotype outcome and 403,195 individuals with morning person outcome (252,287 morning people *vs* 150,908 evening people) were included ^39^. We also evaluated morningness, for which we had information on 345,552 subjects ^36^. Sleep duration phenotypes (continuous and categorical variables) comprised 446,118 ^40^ and 384,317 individuals ^36^. For insomnia two different datasets were used: data from 453,379 subjects reported in the study by Lane *et al*. (2019) ^37^, and data from 386,533 individuals from the Jansen *et al*. (2019) study ^36^. We used only UK Biobank participants’ data due to restrictions in the availability of 23andMe summary statistics for studies that used both ^36,39^. The remaining phenotypes were: ease of getting up (N=385,949), daytime napping (N=386,577), daytime dozing (N=386,548), snoring (N=359,916) and daytime sleepiness (N=45 2,071) ^36,38^. Details on sample and SNP quality control can be found in UKBB documentation ^44^

We also used a GWAS for sleep duration (as a continuous measure) in children ^41^, a result of a meta-analysis of 10,554 children of European ancestry, performed by the EArly Genetics and Life course Epidemiology (EAGLE) Consortium ^43^. Further description of the studies and phenotypes included in this study is given in the Supplementary Material (Table S1).

### Statistical analysis

#### Quality control datasets

As quality control checks, the datasets for ADHD and sleep-related phenotypes were filtered to include only those markers in autosomal chromosomes with (i) an imputation quality score (INFO) score ≥ 0.8, (ii) minor allele frequency (MAF) ≥ 0.01, and (iii) outside the major histocompatibility complex (MHC) region (chr6: 26-33M), according to the information available in each set of summary statistics.

#### Genetic correlation analysis

We performed a cross-trait linkage disequilibrium score regression (LDSC) analysis to estimate the genetic correlation between sleep-related phenotypes and ADHD using the quality-controlled summary statistics of the largest GWAS available for each phenotype. The method involves regressing summary result statistics from millions of genetic variants across the genome on a measure of each variant’s ability to tag other variants locally (i.e. its ‘LD score’) ^45^. LDSC estimates the genetic correlation between two traits (ranging from –1 to 1). For the LD-regression, we used pre-computed LD scores based on European samples from the 1000 Genomes Project (phase 3 version 5) as a reference panel ^46^. The Benjamini and Hochberg False Discovery Rate (BH-FDR) was used to correct for multiple testing (45,46).

#### Gene-based cross-trait/disorder meta-analyses

For the sleep-related phenotypes that were significantly correlated with ADHD in the cross-trait LDSC regression analysis, we performed gene-based cross-trait meta-analysis aiming to uncover putative shared pathways between the phenotypes. For this analysis, we used the same quality-controlled datasets used for the LDSC regression analysis. We used MAGMA software (version v1.07b for Linux) to run a gene-level association analysis to investigate the association of each gene with each phenotype based on p-values reported in summary data ^49^. A conservative Bonferroni correction to account for multiple testing (dividing the significance p-value threshold of 0.05 by the number of genes included) was used. First, variants were mapped to a gene if they were within 10kb upstream or downstream from the gene according to NCBI 37.3 gene definitions. Then, we performed the gene-level association analysis to compute gene p-values for each phenotype, and, thereafter, we ran the cross-trait meta-analyses of gene-level results (ADHD *vs* sleep-related phenotypes). The weighted Stouffer’s Z method was used to combine the Z-scores for each gene across cohorts, with weights set to the square root of the sample size each Z-score is based on (i.e. therefore accounting for the fact that sample sizes vary per single-nucleotide polymorphism (SNP) – and thus per gene – within and between GWAS summary statistics). To identify strongly associated genes in the meta-analysis, we selected genes that showed an increase in their association p-value of at least one order of magnitude compared to the association p-values obtained for each of the individual phenotypes (i.e. ADHD and sleep phenotypes), as has been done previously (Tables S2-S10) ^50^.

#### Gene-set enrichment analyses

To obtain insight into putative biological mechanisms of shared genetic factors found in the previous analyses, we conducted pathway analyses using the MAGMA gene-set implemented in the FUMA 1.3.6 ^51^ online tool (available at http://fuma.ctglab.nl). Enrichment of prioritized genes in biological pathways and functional categories was tested using the hypergeometric test against gene sets obtained from MsigDB ^51,52^. A minimum of 5 genes was considered to characterize a gene set. The coding human genome was used as a reference set, and the BH-FDR correction was used to account for multiple testing.

#### Bidirectional Two Sample Mendelian Randomization (TSMR) analysis

The selection of the instrumental variables (IVs) to run the TSMR analysis was carried out as follows. We clumped the genetic variants available in the summary statistics of the phenotypes: ADHD ^42^, sleep duration phenotypes ^40^, insomnia ^37^, and daytime sleepiness ^38^. We used PLINK 1.9 software to conduct the clumping (r^2^ = 0.05, kb = 500, p1= 5×10^-8^ and p2 = 0.5). There was no SNP associated with sleep duration in children, then we did not included this summary statistics in our TSMR ^41^. For phenotypes that reported the results of a meta-analysis with the 23andMe sample, we opted for keeping the reported top hits to ensure as much statistical power as possible. This criterium was applied for the phenotypes included in the Jansen *et al*. (2019) study (insomnia, napping, snoring, daytime dozing, morningness, ease getting up and sleep duration) as well as in the Jones *et al*. (2019) study (morning person and chronotype as a continuous variable) ^36,39^.

The TSMR analysis was carried out using the R package MR-Base ^53^ available at github.com/MRCIEU/TwoSampleMR). When the selected (index) genetic IV was not available in the outcome GWAS, we replaced it by a proxy variant (R^2^> 0.80) when possible, using as a reference the European population from 1000 Genomes (phase 3 version 5)^46^. Proxies for instruments were identified using the SNP Annotator (SNiPA) v3.2 and LD link tools ^54,55^.

We estimated the effect of sleep-related phenotypes on ADHD (and vice-versa) using the inverse variance weighted (IVW) estimator, which performs a linear regression of the IV-outcome association estimates on the IV-exposure association estimates, weighted by the inverse of the variance of the IV-outcome association estimates. The intercept of this regression is constrained at zero, which corresponds to the assumption that the IVs can only affect the outcome through the exposure or that horizontal pleiotropic effects are balanced ^56^.

As sensitivity analyses, we used other MR methods that are more robust to horizontal pleiotropy than the IVW estimator: MR-Egger regression, penalized weighted median (PWM), and the weighted mode-based estimator (MBE). The MR-Egger method provides a valid test of directional or unbalanced pleiotropy (intercept p-value) and a consistent estimate of the true causal effect ^57^. A Generalised Summary-data-based Mendelian Randomization (GSMR) test was used as an additional sensitivity analysis to detect pleiotropic effects, but was applied only on the datasets that reported the effect allele frequency ^37,40^. We used the platform Complex Trait Genetics Virtual Lab (CTG-VL, available on: https://vl.genoma.io/) to run the statistical package GSMR ^58^. This program implements a method that uses summary data from GWAS to distinguish causal or pleiotropic associations ^59^. We also performed a leave-one-out analysis to identify potentially overly influential IVs by removing one genetic variant at a time and recalculating the IVW estimate.

## Results

### Genetic correlation analysis

The LDSC regression analyses indicated strong positive correlation between ADHD and the following traits: insomnia (Lane *et al.:* r_g_=0.304, se=0.033, P_FDR_=5.8×10^-19^; Jansen *et al.:* r_g_=0.368, se=0.036, P_FDR_= 2.9×10^-23^), daytime sleepiness (Wang *et al.:* r_g_=0.124, se=0.036, P_FDR_=0.001), daytime dozing (Jansen *et al.:* rg=0.296, se=0.058, P_FDR_= 1.1×10^-6^), daytime napping (Jansen: r_g_=0.189, se=0.049, P_FDR_=2.1×10^-4^), snoring (Jansen *et al.:* r_g_=0.166, se=0.036, P_FDR_=8.5×10^-6^), short sleep duration (Dashti *et al.:* r_g_=0.3318, se=0.0366, P_FDR_=6.1×10^-19^) and long sleep duration (Dashti *et al.:* rg=0.384, se=0.046, P_FDR_=3.7×10^-16^). Evidence of a negative correlation between ADHD and sleep duration as a continuous variable ^36^ was observed, which was stronger for the Jansen study ^36^ (r_g_=-0.089, se=0.035, P_FDR_=0.020), than for the Dashti *et al*. study ^40^ (r_g_=-0.067, se=0.038, P_FDR_=0.078), or the Marinelli *et al*. study (r_g_=0.091, se=0.096, P_FDR_ =0.394). We did not observe significant correlation between ADHD and chronotype (Jones *et al.:* r_g_=0.015, se=0.030, P_FDR_=0.607), morningness (Jones *et al.:* r_g_=-0.030, se=0.031, P_FDR_=0.397; Jansen *et al.:* r_g_ =0.045, se=0.029, P_FDR_=0.173), and ease getting up (Jansen *et al.:* rg=0.030, se=0.031, P_FDR_=0.394) phenotypes (Figure 2).

**Figure 2.**
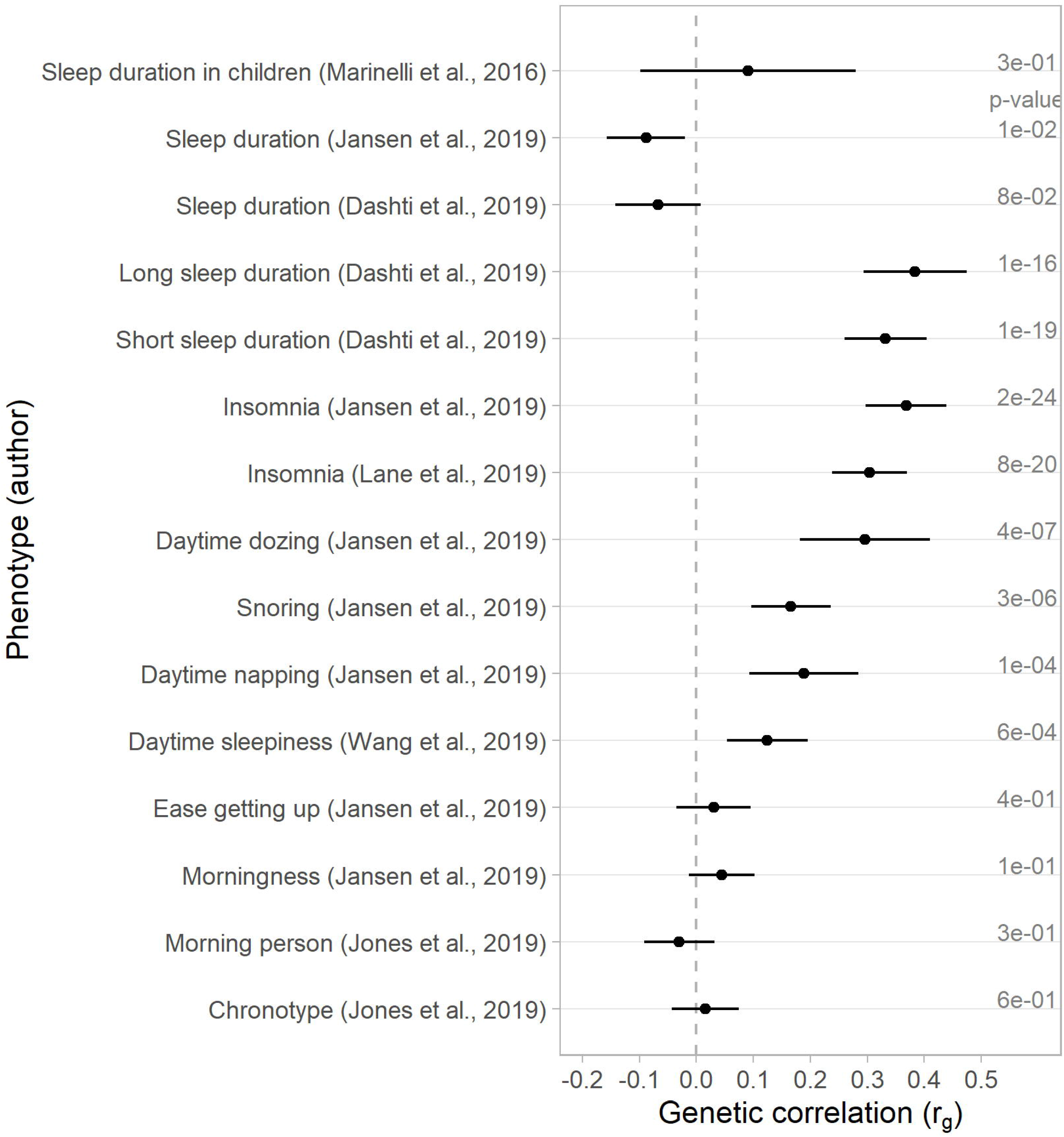
Forest plot depicting the genetic correlations (r_g_) between sleep-related traits and attention deficit/hyperactivity disorder (citation of the summary statistics used). Note: Bars represent 95% confidence intervals

### Gene-based cross-trait meta-analyses and functional enrichment analyses

Regarding the cross-trait meta-analyses, we observed that insomnia had the greatest number of genes shared with ADHD (75 and 44 genes, respectively for the Lane *et al.*., 2019 and Jansen *et al.*., 2019 studies), followed by sleep duration (48 genes), daytime sleepiness (38 genes), snoring (33 genes), short sleep duration (32 genes), long sleep duration (14 genes), and napping (8 genes), whereas only two genes were shared with daytime dozing *(PCDH7; AMMECR1L)*. The gene-set functional enrichment analysis showed significant enrichment at different levels and several shared pathways between phenotypes (Table 1). The genes associated with insomnia and

**Table 1.**
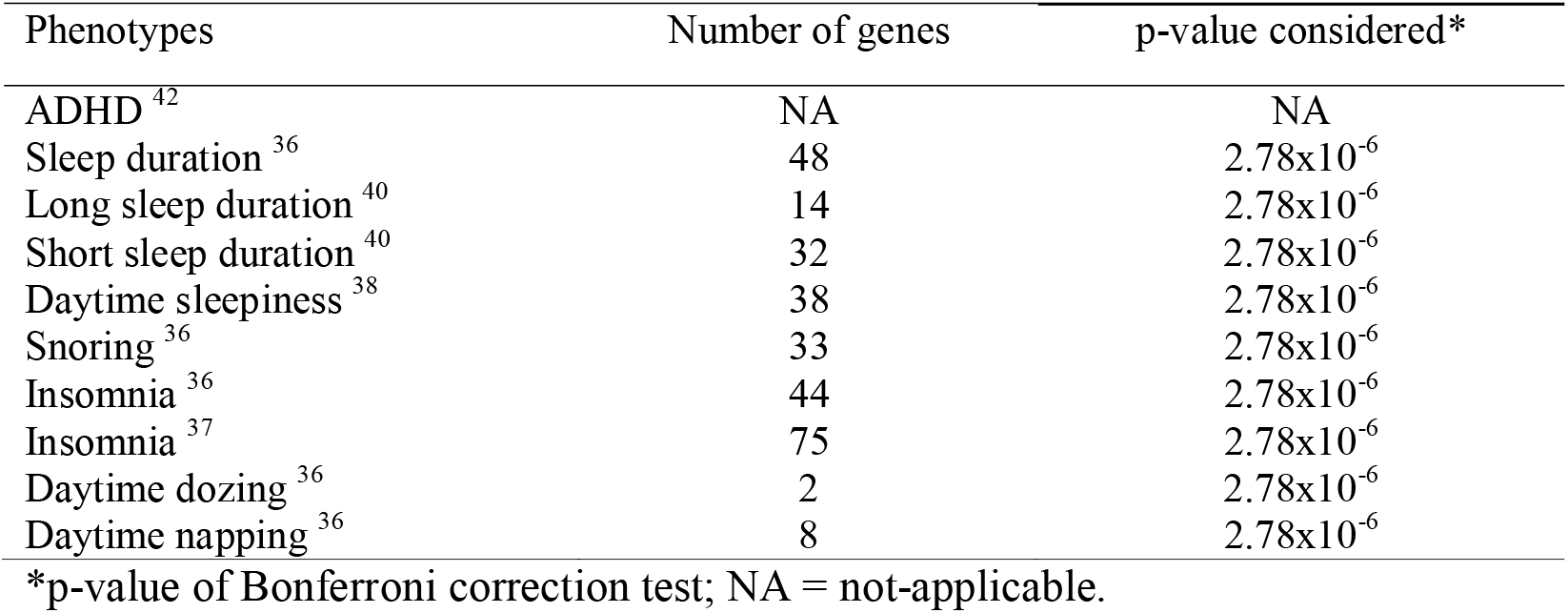
Genes associated with both sleep related traits and Attention Deficit/Hyperactivity Disorder (ADHD) after cross trait meta-analysis

ADHD were enriched in receptor tyrosine kinase signaling (R-HSA-9006934, P_FDR_=3.5×10^-2^), synapse organization (GO:0050808, P_FDR_=1.5×10^-2^) and regulation of synapse structure and activity (GO:0050803, P_FDR_=3.1×10^-2^), and presented a common regulation by *MIR125B* and *MIR125A* (microRNA targets, MsigDB c3, P_FDR_=0.046). Shared genes between daytime sleepiness and ADHD were enriched in synapse (GO:0045202, P_FDR_=6.5×10^-3^) and plasma membrane region (GO:0098590, P_FDR_=6.5×10^-3^; G0:0097060, P_FDR_= 0.016), as well as cell body (GO:0044297 P_FDR_=0.046). They also have a shared gene expression regulation mechanism involving *MIR200B, MIR200C* and *MIR429* (microRNA targets, MsigDB c3, P_FDR_=0.024). Snoring and ADHD shared a group of genes enriched in neurogenesis processes (GO:0022008, P_FDR_=9.1×10^-4^), neuron differentiation (GO:0030182, P_FDR_=7.3×10^-3^), nervous system (GO:0007417, P_FDR_=0.02) and head development (GO:0060322, P_FDR_=0.03). Sleep duration and ADHD shared genes are involved in miRNA regulation and processing, especially those participating in splicing and metabolic processes (microRNA targets). They also shared a common regulatory mechanism including *MIR34A, MIR34C, MIR449, MIR381, MIR204, MIR211, MIR374, MIR27A and MIR27B* (Table 2). For the other phenotypes, no statistically significant functional enrichment was detected.

**Table 2.**
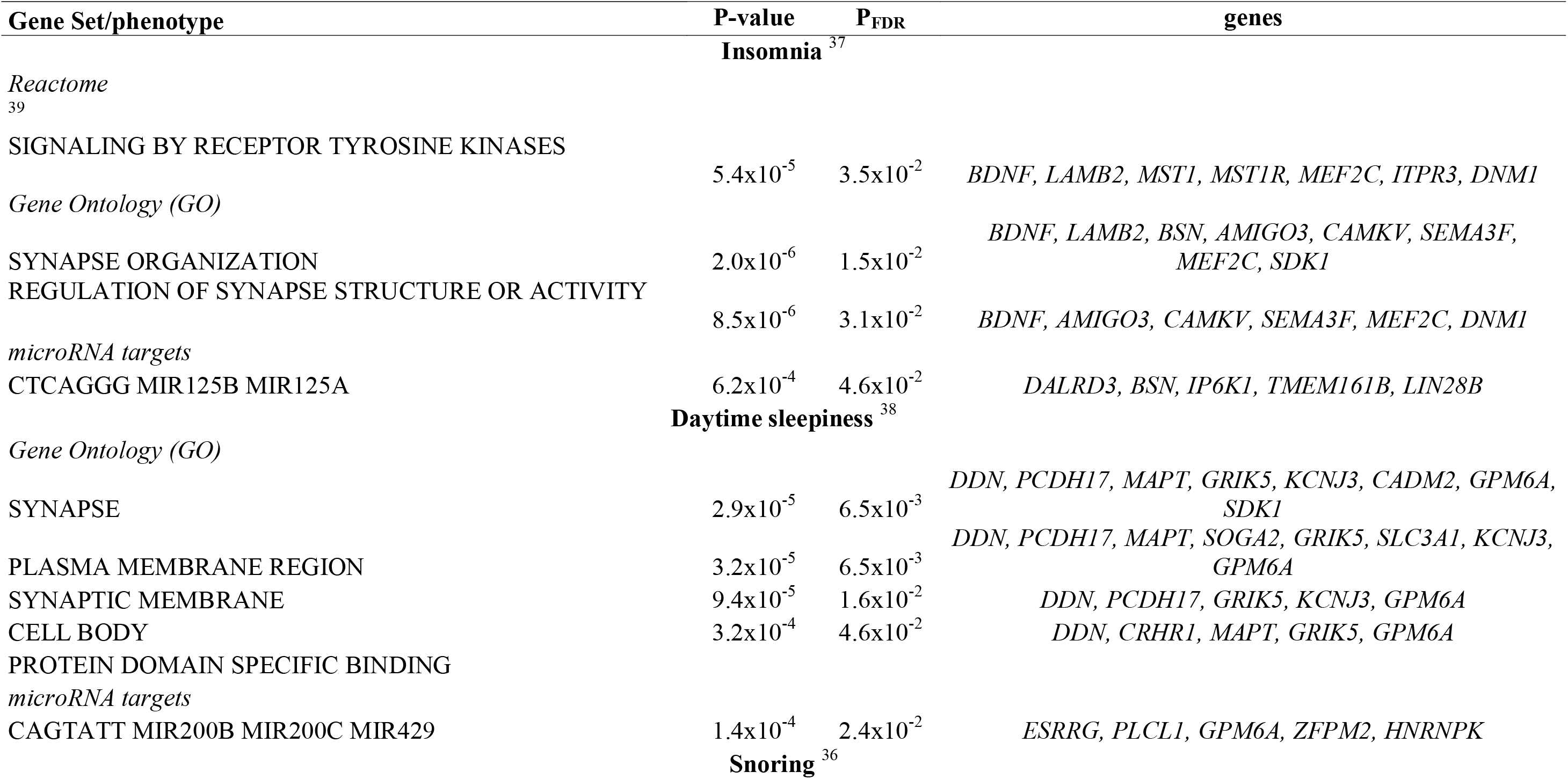
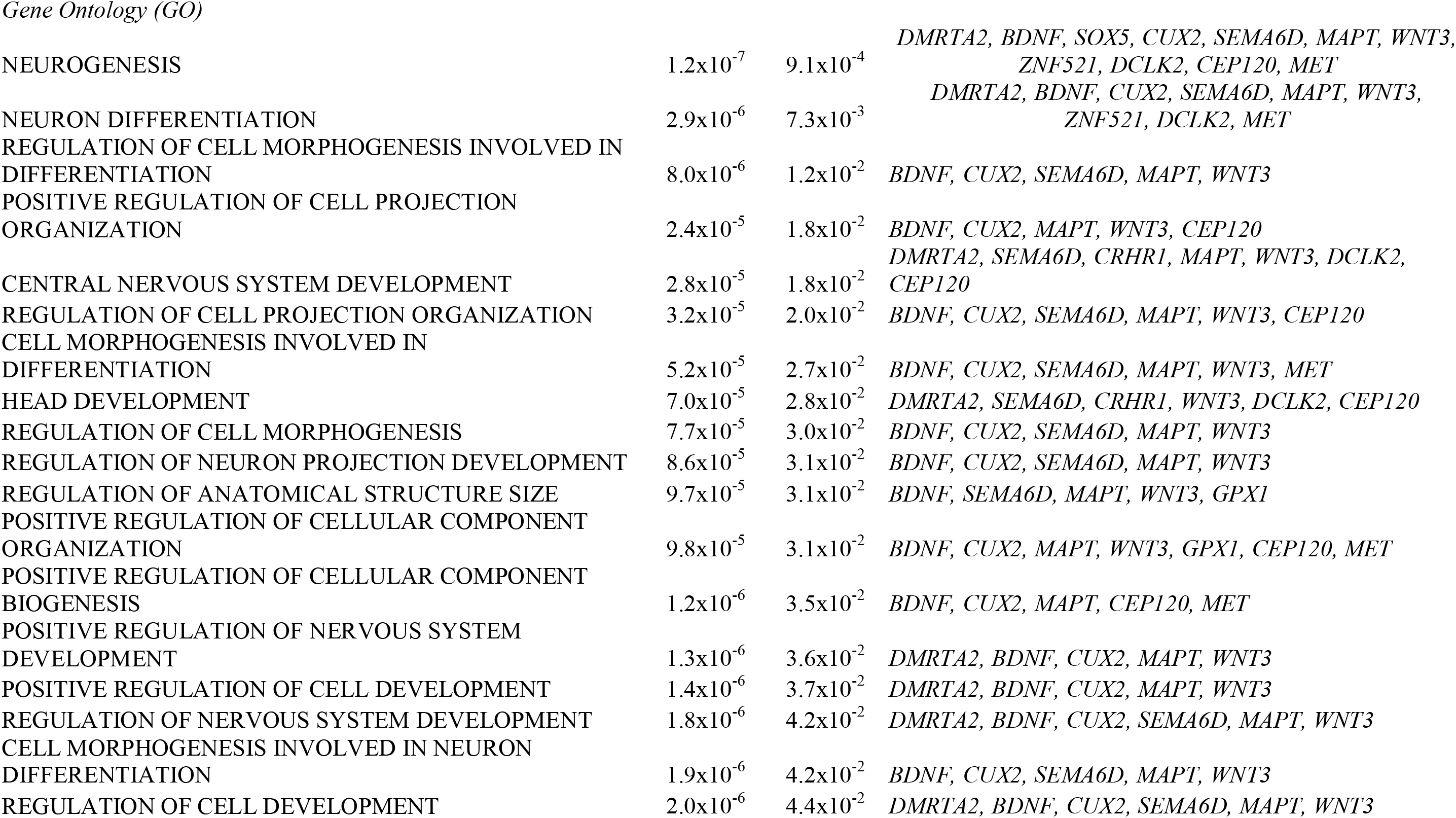
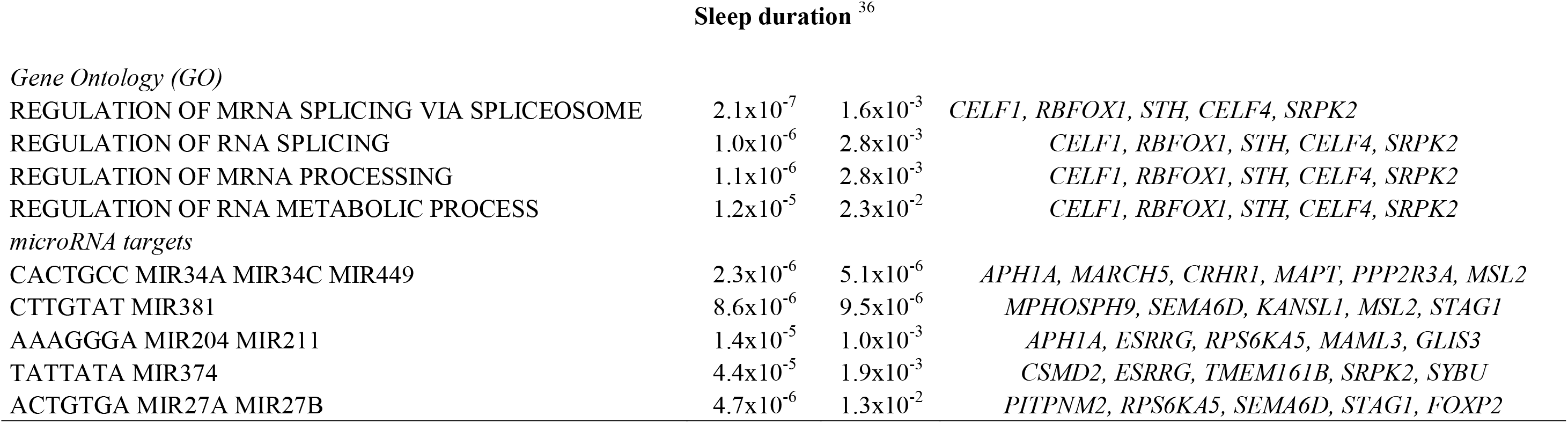
Functional enrichment analyses based on functional annotations of the genes shared by sleep-related phenotypes and Attention Deficit/Hyperactivity Disorder (ADHD) as result of cross-trait metanalyses

### Bidirectional two-sample Mendelian randomization analysis

#### Sleep-related phenotypes as a cause of ADHD

The IVs used in this analysis are presented in Table S11. We found evidence of a causal effect of insomnia (Jansen *et al.:* β = 0.335, SE=0.034, p < 0.001; Lane *et al.:* β = 0.872, SE=0.373, p = 0.019), daytime napping (β = 0.350, SE=0.095, p < 0.001) and short sleep duration (β = 2.032, 0.788, p = 0.010) on ADHD using the IVW estimator. We obtained similar results for the PWM method. However, the results for MR-Egger and the MBE method did not support causality. For short sleep duration the MR Egger coefficient showed inverse direction to IVW and PWM (Figure S1). No evidence of pleiotropy was observed according to the MR Egger intercept test (Table S12) and the GSMR (Table S13) for any of the phenotypes. The leave-one-out analyses showed no disproportionate influence of any specific IV on the pooled IVW-estimated effect (see Figures S1-S12).

#### ADHD as a cause of sleep-related phenotypes

We found that ADHD susceptibility was causally associated with longer sleep duration (β = 0.012, SE=0.003, p < 0.001). In sensitivity analyses, we observed similar results for PWM (β = 0.012, SE=0.004, p = 0.005), but not for MR-Egger and MBE (Table 3). There was no evidence of pleiotropy according to the intercept MR Egger test (Table S12) and the GSMR analyses (Table S13).

Although we did not find evidence of a causal effect of ADHD on chronotype with the IVW method, we detected an inverse association of these phenotypes using PWM (β = -0.049, SE=0.017, p = 0.004), MR Egger (β = -3.150, SE=0.119, p = 0.033) and MBE (β = -0.050, SE=0.020, p = 0.037). The MR Egger intercept test suggested the presence of pleiotropy acting on this association (intercept = 0.026, SE = 0.01, p = 0.043) (Table S12). We also found a negative effect of ADHD on morningness (PWM β = -0.039, SE=0.015, p = 0.007), as well as of ADHD on being a morning person (MR Egger β = -0.509, SE=0.204, p = 0.042, MBE β = -0. 078, SE=0.032, p = 0.015) in the sensitivity analyses. (Table 3). In these cases, the evidence of pleiotropy was weaker (Table S12). The leave-one-out analyses showed no disproportionate influence of any specific IV on the pooled IVW-estimated effect for most of the phenotypes (see Figure S13-S22). However, for morning person (Figure S23) and chronotype (Figure S24) we observed that when we removed rs10262192, the IVW association become significant.

**Table 3.**
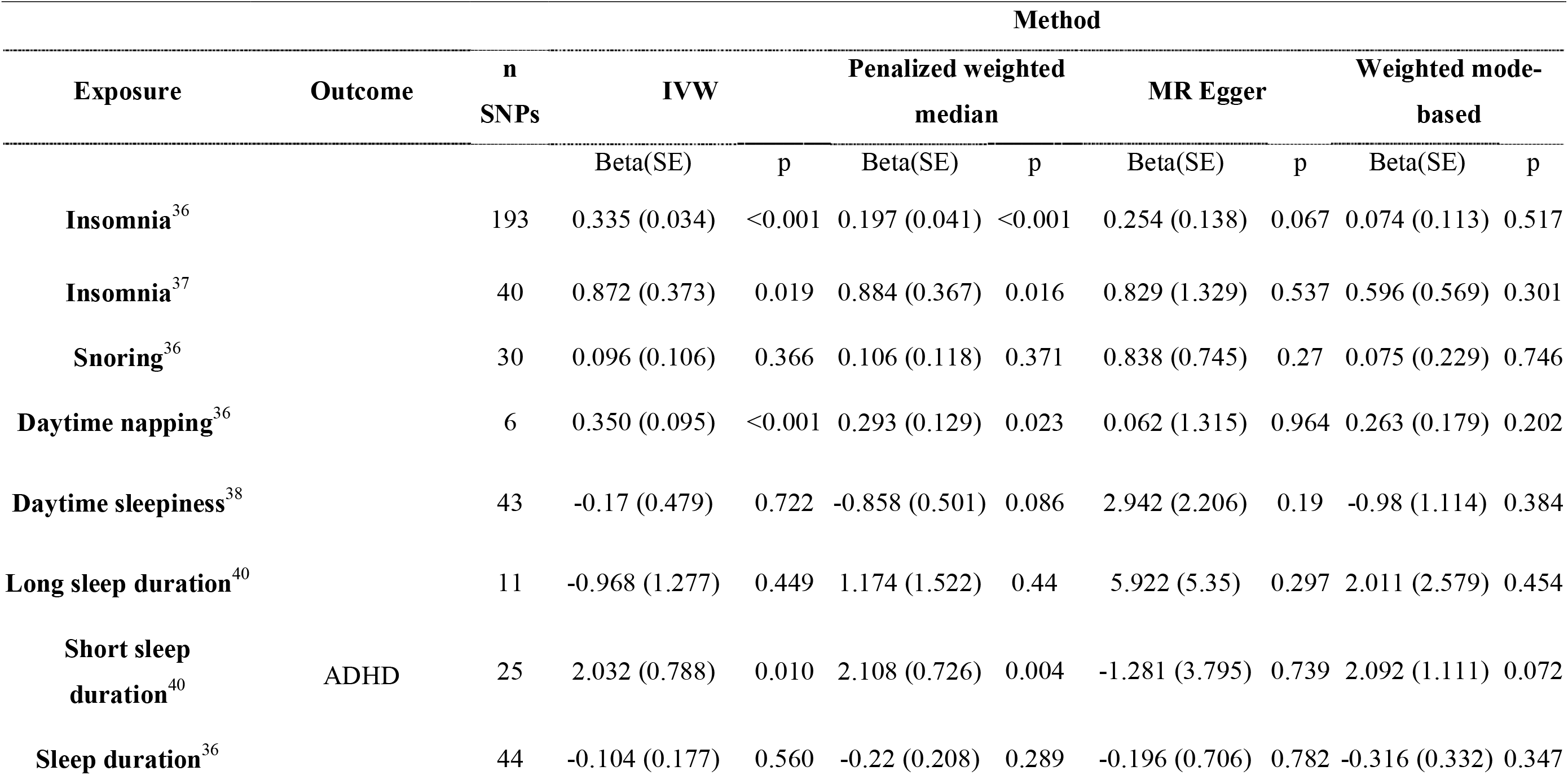
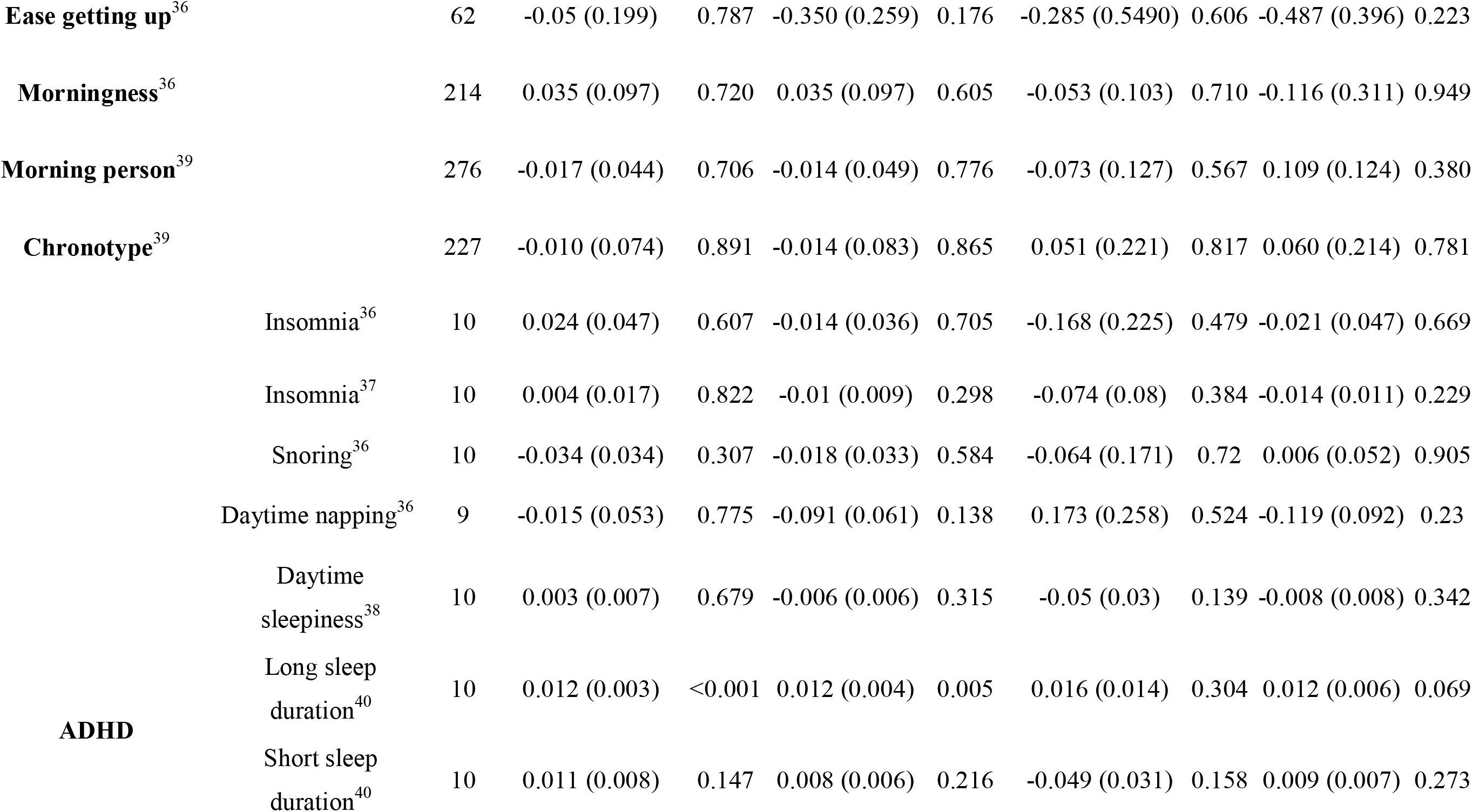
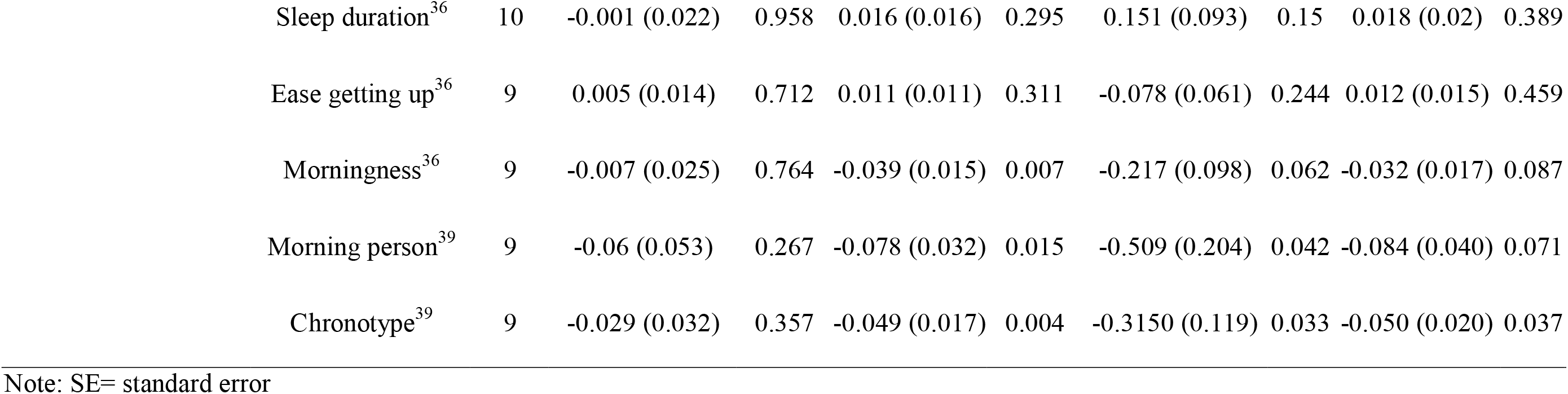
Bidirectional two-sample Mendelian randomization analyses estimating the causal effect of sleep-related phenotypes on Attention Deficit/Hyperactivity Disorder (ADHD) risk and vice-versa

## Discussion

In this study we showed that several sleep-related phenotypes, mainly sleep disturbances, and ADHD are genetically correlated, having potentially shared mechanisms, mostly involving neurodevelopment, neurogenesis, and synaptic organization. The results were less consistent for phenotypes related to sleep duration, and we did not find evidence of shared genetics or mechanisms with chronotype phenotypes. We also found evidence of a causal effect of insomnia, daytime napping, and short sleep duration on ADHD, and of ADHD on long sleep duration and morningness.

Our results suggest that sleep disturbances, sleep duration and ADHD have a shared inheritance, given the strong genetic correlations we detected. Similarly, earlier studies have reported an association between polymorphisms in core clock regulation genes and ADHD using candidate-gene study designs ^21-25^. Also, the expression of different clock genes has been associated with ADHD in human and animal models ^1-4^. Our results are in accordance with studies showing that both ADHD and sleep disturbances are linked to aberrant brain development ^34,60-63^, display neuroanatomical similarities in subcortical and cortical areas ^64,65^ as well as overlapping cortico-strial circuitry ^66-69^. At the molecular level we found evidence of a shared mechanism between ADHD and sleep disturbance, specially insomnia and daytime sleepiness, and between ADHD and sleep duration. The group of genes that we found as being shared by sleep deprivation-related phenotypes (insomnia, daytime sleepiness and sleep duration) and ADHD is associated with synapse structure, activity, organization and regulation. We also uncovered genes involved in the regulation of brain? anatomical structure and size and neurodevelopment function amongst those shared between snoring and ADHD. A neurobiological substrate linking ADHD and sleep disturbance, specially dyssomnia, was recently reported, showing neuroanatomical impairment using neuroimage data and suggesting a role of genes related to dopamine signaling and the circadian cycle ^34^, which corroborates our findings.

We found a group of miRNAs shared by sleep deprivation-related phenotypes and ADHD that are involved in posttranscriptional gene silencing and are mostly expressed in brain regions, especially in the cortex and cerebellum *(MIR204, MIR27B, MIR125A, MIR429)*. These results point towards a role of epigenetics on the relation between both traits. The effect of sleep deprivation on the transcriptome and methylome has previously been studied using animal models and human subjects ^70-75^. Whilst other studies indicate that sleep deprivation induces notable changes in the brain transcriptome of rats, affecting protein synthesis, synaptic plasticity, and metabolism ^72,76^.

So far there is no consensus about the direction of the sleep-ADHD association, despite evidence showing that sleep deprivation and sleep disturbances have known cognitive emotional and behavioral effects that mimic ADHD ^3,26,31^, and that early sleep disturbances are associated with an increased risk of ADHD later in life ^28,29,31,33,77^. The TSMR analyses we carried out showed a causal effect of insomnia, daytime napping, and short sleep duration on ADHD. Interestingly, our findings corroborate the neuropsychological-biology theory of the prominent role of sleep, especially sleep deprivation as a predisposing factor to ADHD, probably through executive function deficits (for a review see references^3,4,11,78,79^). Considering that sleep disturbances in children worsen executive functions, and this is an important characteristic of ADHD patients, we hypothesize that sleep problems may cause biochemical imbalances in executive function, and therefore predispose the individual to ADHD ^11,66,69,79,80^.

On the other hand, we found a causal effect of ADHD on long sleep duration, and on morning chronotype after accounting for pleiotropy. Longer sleep usually reflects improved, higher quality sleep, because of potentially increased sleep efficiency. However, it may also bring about qualitatively poorer sleep, or greater sleep need, given the association with a larger number of sleep bouts and increased daytime napping, in addition to being a risk factor for cognitive dysfunction and other negative health outcomes^40,81,82^. It is possible that the ADHD symptomatology would provoke mental distress which may lead to longer hours of sleep. The causal effect of chronotype on mental health has been already reported ^39^. Here, we found that ADHD may be a causal factor for manifesting an evening chronotype, a direction consistent with that reported in observational studies ^3^. Therefore, although our findings suggest a bidirectional association between sleep and ADHD, as reported before ^33,35^, additional studies should be undertaken to explore this relationship further.

Several observational studies have reported that sleep disturbances and short sleep duration are likely predisposing to ADHD^32,35,35^, whilst others have uncovered effects in the opposite direction^32,34,35^. However, inferring causality in observational studies is problematic due to unmeasured confounding, reverse causality, and other unknown biases ^84,85^. MR is a method to infer causality which uses genetic variants robustly associated with an exposure variable as instruments to test causal effects on an outcome variable. The advantages of using genetic variants underlying a trait include that they are unlikely to be associated with confounders, and that genotypes are randomly allocated and set at conception, thus considerably reducing the risk of bias introduced by confounding and reverse causation ^84,86,87^. Using TSMR our study provided stronger evidence of a causal effect of sleep deprivation-related phenotypes on ADHD, as well as an effect of ADHD on long sleep duration and evening preferences.

Our results should be interpreted considering some limitations. The first concerns the potentially different genetic architecture of sleep duration between children and adults ^40^. To address this, we added to our study the only dataset, that we were aware of, that included sleep information in children ^41^. Given the relatively small sample size of this study we were not able to make further inferences about differences across the lifetime, therefore the TSMR analysis should be interpreted with caution concerning the temporality of the phenotypes. Second, our results are based for the most part on UKBB data information. Different datasets are desirable to test the consistency of the findings. However, the scarcity of studies examining sleep and/or ADHD makes it difficult to replicate our results elsewhere. Third, the sleep characteristics used as outcome in the GWAS included in this study were self-reported, which may have led to response bias. However, it is important to highlight that this is an appropriate epidemiology data collection approach for large samples due to logistic and resource availability issues, and that self-reported sleep duration was consistent with accelerometer-estimated sleep duration in a large subsample of the UKBB ^40^. Fourth, most of the GWAS used in our study did not exclude individuals taking sleep (Table S1) or ADHD medication. Sleep medication may have led to sleep phenocopies whereas ADHD stimulant and non-stimulant medication may affect sleep behavior and clock genes at a molecular level ^13,88-90^. Thus, further studies incorporating medication intake are needed. Finally, pleiotropy is an important factor that would violate the exclusion-restriction assumption of Mendelian randomization ^57,84^. We implemented the MR-Egger regression test and GSMR analysis to address this limitation. No evidence of pleiotropy was observed with both tests for most of the analyses. However, a pleiotropic effect was detected on the association of ADHD with chronotype phenotypes.

To our knowledge, this is the first study to investigate the shared genetics and molecular mechanisms between ADHD and sleep-related traits, and their causal relationship, using a genomic approach. Our findings support a positive genomic correlation between ADHD and insomnia, daytime dozing, daytime sleepiness, snoring, napping, and short and long sleep duration, and postulate an important role of neurobiological functions, gene expression and protein synthesis. The causal analysis showed that sleep deprivation-related phenotypes are likely to increase the risk for ADHD, although ADHD cases may be longer sleepers and display an evening chronotype. Therefore, our results suggest that sleep traits and ADHD may be linked by a common genetic background and molecular mechanism as along with a causal relationship.

These findings provide insights into ADHD diagnosis and potential treatment and indicate that sleep deprivation-related phenotypes may act as important ADHD predictors or early markers.

## Data Availability

URLs. UK Biobank website, http://www.ukbiobank.ac.uk/
Functional Mapping and Annotation (FUMA) software, http://fuma.ctglab.nl/
Multi-marker Analysis of Genomic Annotation (MAGMA) software, http://ctg.cncr.nl/software/magma
LD Score regression software, https://github.com/bulik/ldsc
LD Hub (GWAS summary statistics), http://ldsc.broadinstitute.org/
LD scores, https://data.broadinstitute.org/alkesgroup/LDSCORE/; GeneCards, http://www.genecards.org/; Psychiatric Genomics Consortium (GWAS summary statistics), http://www.med.unc.edu/pgc/results-and-downloads; iPSYCH, http://ipsych.au.dk/downloads/; MSigDB curated gene set database, http://software.broadinstitute.org/gsea/msigdb/collections.jsp; GSMR software, http://cnsgenomics.com/software/gsmr/; R package MR-Base, github.com/MRCIEU/TwoSampleMR; CTG-VL, https://vl.genoma.io/.

http://www.ukbiobank.ac.uk/

http://github.com/MRCIEU/TwoSampleMR

http://ipsych.au.dk/downloads/

http://www.med.unc.edu/pgc/results-and-downloads

## Acknowledgment

We acknowledge with gratitude the financial support received for this work. This study was financed in part by the Coordenação de Aperfeiçoamento de Pessoal de Nível Superior— Brasil (CAPES)—Finance Code 001, and by Conselho Nacional de Desenvolvimento Científico e Tecnológico (CNPq, processo no. 427524/2018-0). MXC, AM and LTR were supported by Conselho Nacional de Desenvolvimento Científico e Tecnológico (CNPq). CB was supported by a Universidade de São Paulo/ Coordenação de Aperfeiçoamento de Pessoal de Nível Superior fellowship (processo no. 88887.160006/2017-00). TMS has received support by Coordenação de Aperfeiçoamento de Pessoal de Nível Superior—Brasil (CAPES). Research projects conducted by MHH have been supported by CNPq. JPG has received support by the Research Support Foundation of Rio Grande do Sul—FAPERGS (Fundação de Amparo à Pesquisa do Estado do Rio Grande do Sul). We would also like to thank Guillermo Reales and Christian Loret de Mola for the support on data managing and Angélica Salatino-Oliveira for reviewing our final manuscript.We acknowledge with gratitude the financial support received for this work. We would also like to thank Guillermo Reales and Christian Loret de Mola for the support on data managing and Angélica Salatino-Oliveira for reviewing our final manuscript.

